# Hemodynamic profiles by non-invasive monitoring of cardiac index and vascular tone in acute heart failure patients in the emergency department: external validation and clinical outcomes

**DOI:** 10.1101/2021.11.18.21266430

**Authors:** Nicholas E Harrison, Sarah Meram, Xiangrui Li, Patrick Medado, Morgan B White, Sarah Henry, Sushane Gupta, Dongxiao Zhu, Peter S Pang, Phillip Levy

## Abstract

**Background:** Non-invasive finger-cuff monitors measuring cardiac index and vascular tone (SVRI) classify emergency department (ED) patients with acute heart failure (AHF) into three otherwise-indistinguishable subgroups. Our goals were to validate these “hemodynamic profiles” in an external cohort and assess their association with clinical outcomes.

**Methods:** AHF patients (n=257) from five EDs were prospectively enrolled in the validation cohort (VC). Cardiac index and SVRI were measured with a ClearSight finger-cuff monitor (formerly NexFin, Edwards Lifesciences) as in a previous study (derivation cohort, DC, n=127). A control cohort (CC, n=127) of ED patients with sepsis was drawn from the same study as the DC. K-means cluster analysis previously derived two-dimensional (cardiac index and SVRI) hemodynamic profiles in the DC and CC (k=3 profiles each). The VC was subgrouped *de novo* into three analogous profiles by unsupervised K-means consensus clustering. PERMANOVA tested whether VC profiles 1-3 differed from profiles 1-3 in the DC and CC, by multivariate group composition of cardiac index and vascular tone.

Profiles in the VC were compared by a primary outcome of 90-day mortality and a 30-day ranked composite secondary outcome (death, mechanical cardiac support, intubation, new/emergent dialysis, coronary intervention/surgery) as time-to-event (survival analysis) and binary events (odds ratio, OR). Descriptive statistics were used to compare profiles by two validated risk scores for the primary outcome, and one validated score for the secondary outcome.

**Results:** The VC had median age 60 years (interquartile range {49-67}), and was 45% (n=116) female. Multivariate profile composition by cardiac index and vascular tone differed significantly between VC profiles 1-3 and CC profiles 1-3 (p=0.001, R^2^=0.159). A difference was not detected between profiles in the VC vs. the DC (p=0.59, R^2^=0.016).

VC profile 3 had worse 90-day survival than profiles 1 or 2 (HR = 4.8, 95%CI 1.4-17.1). The ranked secondary outcome was more likely in profile 1 (OR = 10.0, 1.2-81.2) and profile 3 (12.8, 1.7-97.9) compared to profile 2. Diabetes prevalence and blood urea nitrogen were lower in the high-risk profile 3 (p<0.05). No significant differences between profiles were observed for other clinical variables or the 3 clinical risk scores.

**Conclusions:** Hemodynamic profiles in ED patients with AHF, by non-invasive finger-cuff monitoring of cardiac index and vascular tone, were replicated *de novo* in an external cohort. Profiles showed significantly different risks of clinically-important adverse patient outcomes.

## Background

Acute heart failure (AHF) accounts for 1 million emergency department (ED) visits annually in the United States(US), 80% of which result in hospital admission^1,2^. AHF 30-day mortality overall (8-10%^3^) greatly exceeds the threshold of typical emergency physician (EP) risk tolerance (0.5-1%)^4^, and neither EP gestalt for AHF mortality risk^5,6^ nor clinical decision rules (CDRs) yet provide predictive value sufficient^1,7^ to meet such low risk thresholds. Consequently, half of ED to hospital admissions for AHF involve low-risk patients for whom admission may not be neccessary^1,2,8-10^, and recent Society for Academic Emergency Medicine (SAEM) and Heart Failure Society of America(HFSA) guidelines^1^ stress the importance of developing new AHF risk markers in the ED. A particular need exists for novel markers which identify low-risk AHF presentations by way of capturing the high-degree of physiologic and clinical heterogeneity between AHF patients, given relatively more established predictors of high-risk^1,11^ and the high baseline ED admission rate.

Classification by hemodynamic profile is one of the oldest approaches to subgrouping the high clinical heterogeneity present among AHF patients, given that hemodynamic derangements are critical defining features of AHF pathophysiology. Hemodynamic parameters like cardiac index, vascular tone (systemic vascular resistance index {SVRI}), heart rate, blood pressure (BP), and others reflect some of the greatest physiologic heterogeneity among AHF patients^12-18^, and consideration of heart rate and BP are prominent features of contemporary ED AHF evaluation^14^. Cardiac index and vascular tone play an outsized role in AHF pathophysiology^14,15^, yet are not generally able to be assessed in the ED. Gold standard measurement by pulmonary artery catheterization (PAC) requires specialist expertise, is highly invasive, and is employed in only 1% of contemporary AHF hospitalizations^19^. The Forrester classification of “wet-dry/warm-cold” on physical exam^20,21^ is a non-invasive method for assessing cardiac index and vascular tone in AHF^1,22,23^, but limited in clinical utility given a subjective nature and poor reliability, with interrater agreement of just 64% (kappa=0.28) in ED patients^24^.

Recently, a non-invasive monitor providing continuous estimation of cardiac index and vascular tone^18^ was described in an ED-based retrospective study of the PREMIUM (Prognostic Hemodynamic Profiling in the Acutely Ill Emergency Department Patient) registry by Nowak et. al. ClearSight (formerly “NexFin”, as it was known in this prior study; Edwards Lifesciences, Irvine, California) is an FDA-approved finger-cuff monitor which measures continuous blood pressures and pulse rates at both the radial and digital arteries. Finger-cuff monitors are attractive for ED profiling of patients by cardiac index and vascular tone, because they are non-invasive like the Forrester classification, yet provide the clinician with reproducible, continuous, and objective measurements like a PAC. Nowak et al. in turn derived 2-dimensional hemodynamic profiles by cardiac index and vascular tone (SVRI) from the finger cuff measurements in these ED AHF patients. Profiling by these two hemodynamic variables is the same physiological construct underlying the Forrester classification, but with objective measurements guiding classification rather than highly-subjective^24^ physical examination. Namely, profiling by cardiac index and vascular tone reflects that the ventricular-vascular relationship is naturally discordant, since maintenance of blood pressure requires any decrease in cardiac index to be buffered by an equal and opposite increase in vascular tone and visa-versa (i.e. Mean arterial BP = cardiac index x SVRI)^14^. Importantly, disruption of the ideal ventricular-vascular relationship, such as by nitrate metabolism, myocardial changes, neurohormonal effects, and other factors, is a key component of HF physiology^5,11,15,16,25-30^. Moreover, this construct acknowledges that AHF patients are highly heterogenous in the cardiac index vs. vascular tone relationship, as demonstrated in Nowak et al.’s study of ED patients^18^.

Three distinct ED AHF hemodynamic profiles by CI and vascular tone were described by Nowak et al. which, critically, were not otherwise identifiable by clinical characteristics^18^. This 3-profile system observed reflected a surprisingly large degree of previously uncaptured physiologic heterogeneity^18^, spanning very low and very high values in both the cardiac index and vascular tone dimensions. While potentially promising, the prior study was not powered to detect prognostic difference between profiles, and thus clinical significance of these groups is unknown. Additionally, the use of finger-cuff monitors in non-invasive ED AHF hemodynamic monitoring is novel, and the ability to reproduce this 3-profile classification has not been shown in an external sample.

In the current study, we sought to validate the hemodynamic profiles derived in the prior study, and to assess whether they correlate to adverse outcomes. We hypothesized that 1.In De novo cluster analyses of a new prospective sample of AHF patients (validation cohort, VC) would produce cardiac index/vascular tone hemodynamic profiles matching the derivation cohort(DC)^18^; 2. To demonstrate the uniqueness of the profiles to AHF, VC profiles would not match profiling of patients in PREMIUM with sepsis (control cohort, CC), and 3. Between-profile differences in the VC would exist for both 90-day all-cause mortality (primary outcome) and a 30-day ranked composite of adverse events.^31,32^ (secondary outcome).

## Methods

CLEAR-AHF was a multicenter prospective observational study approved as minimal-risk research by the Wayne State University (WSU) and Indiana University (IU) institutional reviews boards. The primary aim of the study was to create a multi-institutional registry of hemodynamic data in ED AHF patients, given the lack of other methods for measuring important hemodynamic parameters like cardiac index and SVRI (vascular tone) in the emergency department. A secondary aim was to validate the hemodynamic profiles derived in the pilot study^18^ and determine whether these profiles were associated with clinically-important patient outcomes. Written consent was obtained from all participants. This manuscript was written to comply with the STROBE guidelines for cohort studies^33^. The ClearSight monitor’s manufacturer, Edwards Life Sciences, provided funding for this investigator-initiated study including research assistant time, the monitors used in the study, and salary support for one of the site principal investigators (PL). Edwards was not involved in the design, planning, conduct, analysis, or written report of this investigation.

### Study setting and participants

Patients ≥18 years old presenting to the ED were screened for enrollment 24 hours per day at five EDs in the US. Annual ED volumes range from approximately 80,000 - 100,000 patient visits per site. Enrollment occurred from July 2017 - March 2019.

Included patients had EP suspicion of AHF and at least one of the following: dyspnea at rest or exertion, signs of AHF on chest radiograph (CXR), and/or NT-proBNP>300 pg/ml. Patients with dyspnea primarily due to other causes (by EP diagnosis), temperature >38.5 °C or suspected sepsis, acute ST-elevation myocardial infarction (STEMI), pregnant women, prisoners, and those without an ejection fraction (EF) recorded within 12 months, were excluded. Two study authors with extensive AHF research experience (PP and PL), blinded to one another, performed case review to adjudicate the ED diagnosis of AHF. Disagreements were decided by discussion, and patients without diagnostically-adjudicated AHF were excluded.

### Study protocol

Patients were fitted with a ClearSight finger-cuff monitor and continuous measurements recorded for 3-6 hours. Manufacturer reference range values for cardiac index and SVRI are 2.5-4.0 L/min/m2 and 1970-2390 dynes-sec/cm5/m2, respectively. Clinicians and patients were blinded to device measurements by an opaque sheet on the monitor screen to prevent bias in management decisions. Patients were excluded if they could not begin monitoring in the ED. The median time from initial IV loop diuretic to initiation of hemodynamic monitoring in the sample was 98 minutes (IQR: 29-167), and the first recorded value for cardiac index and SVRI were used for the analysis.

Research assistants used standardized data sheets to record patient demographics, medications, medical history, vital signs, clinical tests, ED treatments, ED disposition, and hospital course. Data were obtained through patient phone interview, the electronic medical record (EMR), and the reports of treating physicians. Data were recorded in REDCap (Research Electronic Data Capture; http://project-redcap.org/).

### Measures

#### Primary Measure

The primary measure of interest was each patient’s 3-level categorical hemodynamic profile, as derived in the DC^18^. Each profile is a subgroup of the multivariate distribution in two dimensions: cardiac index and vascular tone (SVRI). The first available cardiac index and vascular tone, obtained simultaneously, was used for profiling (see Analysis, Validation Parts 1-2). Finger-cuff hemodynamic monitors have >90% correlation to invasive arterial blood pressure monitoring^34^, and estimate invasive cardiac index with a ∼30% margin of error^18,35^. While the level of accuracy of this estimate does not imply interchangeability with invasive hemodynamic monitors^27^, it is nevertheless accurate enough to be useful for initial assessment before an invasive monitoring can begin in the intensive care unit^27^. Since invasive hemodynamic monitoring is only performed in 1% of contemporary AHF hospitalizations^19^ and is outside the scope of practice of many EPs, a non-invasive estimate such as from a finger-cuff monitor is the only feasible measure for cardiac index in the ED. For more details on finger-cuff hemodynamic monitors, see the publication for the DC^18^ and other prior literature^27^.

Cluster analysis methods to subgroup patients by cardiac index and vascular tone into one of three hemodynamic profiles are described below for the VC (see Validation Part 1) and in the prior publication^18^ for the DC. The CC hemodynamic profiles were obtained in an unpublished analysis by the same methods and at the same time as the DC^18^, from a concurrently enrolled group of septic patients in the same registry (PREMIUM) as the DC’s AHF patients.

Hemodynamic profiling by cardiac index and vascular tone was performed *de novo* in the VC (see Validation Part 1), to test if profiling of the VC replicated the profiles of AHF patients in the DC^18^ (hypothesis 1) and differed from the profiles of non-AHF patients in the CC (hypothesis 2). For consistency, profile numbering 1 through 3 in the VC was set to match the corresponding profiles 1-3 in the DC and CC.

#### Secondary Measures

Other measures included numerous clinical variables: demographics, vital signs, laboratory tests performed in the ED, chest x-ray (CXR) and electrocardiogram (ECG) findings. Three validated CDRs for AHF-risk stratification in the ED were also calculated as well to place our study in context. The Emergency Heart Failure Mortality Risk Grade (EHMRG) and Get With The Guidelines HF Risk Score (GWTG-HF) have were derived^5,36^ and externally validated^37-39^ to predict short-term AHF mortality. The STRATIFY risk score was derived^1^ and validated^32^ in a US ED population to predict a 30-day ranked composite of clinically-important AHF adverse events (study secondary outcome, least to most severe): 1. invasive cardiac procedure or acute coronary syndrome (IP/ACS), 2. new or emergent dialysis (NED), 3. intubation, 4. mechanical cardiac support or transplant (MCS/T), 5. death or cardiopulmonary resuscitation (D/CPR). The GWTG-HF risk score has been shown to have both short-term^36^ and long-term^38,39^ prognostic value for AHF mortality. EHMRG was previously demonstrated to outperform EP gestalt^5,37^ for short-term AHF mortality.

### Outcomes

The primary clinical outcome was mortality within 90 days of ED presentation. The secondary outcome was the hierarchical 30-day composite used in STRATIFY, described above (Secondary Measures). All events in the secondary outcome hierarchy were recorded for each patient. Multiple occurrences of the same event were treated as a single event for analysis purposes. The primary and secondary outcomes were recorded as both days-to-event (survival) and binary variables.

Study coordinators collected outcome information by EMR follow-up and telephone interview at 30 days, 90 days, 180 days, and 1 year. Both institutions have large hospital networks (4 hospitals WSU,14 hospitals IU) sharing EMR data with inpatient and outpatient services. Both also participate in state-wide healthcare information exchanges (HIEs). HIE data was used to augment patient telephone follow-up of adverse events occurring at outside hospital systems. Follow-up through the HIEs, telephone interviews, and local EMRs resulted in 100% confirmation of survival vs. death at 90 days. Records for each outcome were independently queried by two or more data abstractors blinded to the analysis.

### Statistical Analysis

All analyses were conducted in the R statistical programming language (http://www.r-project.org, The R Foundation, v3.6.1). Cluster analysis for the VC was performed in the ConsensusClusterPlus R package^40^. Fully-runnable code and deidentified minimal data sets are included in Supplement S4.

#### Validation part 1: de novo identification of hemodynamic profiles in the validation cohort

We performed cluster analysis of the VC *de novo* to ensure that the cluster algorithm assignment of hemodynamic profiles to VC patients was naïve to all patient data besides cardiac index and vascular tone, and naïve to the data and profiling in the DC^18^. We reasoned that this would help test the criterion validity of the marker of interest. First, if hemodynamic profile on the finger-cuff device represents a reproducible feature of AHF patient physiology, then profiling in the DC and VC should not appreciably differ by multivariate distribution of cardiac index and vascular tone. If they did differ, it would suggest that profiling of the DC represented random statistical noise and/or overfitting of the data rather than a reproducible physiologic feature. Second, we hypothesized the VC profiles would differ in cardiac index and vascular tone from the profiling of ED patients with sepsis in the CC. Namely, if the hemodynamic profiles in the VC represent a construct specific to AHF hemodynamics, the VC profiles should be distinguishable from 3-category profiling of a condition with markedly different hemodynamics like sepsis. Septic patients in the CC were enrolled at the same time and at the same centers as the AHF patients in the DC, as part of the PREMIUM registry.

#### Validation part 2: Unsupervised machine learning cluster analysis method for profiling of the validation cohort

Standard K-means clustering was used in the DC to derive the original hemodynamic profiles^18^ but has two major weaknesses: 1. the data analyst must decide before clustering how many clusters (k) to subgroup the data by, and 2. clustering of the DC was performed without internal validation (i.e. using the entire cohort). Both weaknesses could potentially lead to overfitting of each class/profile to the data set. For the *de novo* profiling of the VC we used a machine learning tool called consensus clustering^40^ to identify two-dimensional patient clusters. As in standard k-means clustering, the input is multivariate data (cardiac index and vascular tone) for each individual in the study. In consensus clustering there is no assumption of the ideal number of clusters, and internal validation to assess cluster stability is performed unsupervised through random resampling and introduction of random data perturbations^40^. The result a reduction in the chance of spurious class discovery due to overfitting of the dataset and less reliance on analyst-provided assumptions. Supplement S1 gives further methodological details on consensus clustering and how it differs from k-means. While the DC study’s authors subjectively chose k=3 profiles for patient classification, we allowed the learning machine to split the data into anywhere between 1-10 unique hemodynamic profiles. Nonetheless, 3-4 groups maximized the internally-validated consensus scoring based on examining consensus score dendrograms (supplement S2) and the elbow plot (supplement S3) of change in cumulative distribution function for each subsequent level k 1-10.

Comparisons of VC profiling with the DC and CC profiles were first made qualitatively by superimposing scatter plots (cardiac index vs. vascular tone) of the cohorts, along with their hemodynamic profiles. Quantitative analyses were performed by PERMANOVA, in which VC profiling was compared to the DC and CC profiles for multivariable similarity of group composition by cardiac index and vascular tone. Cardiac index and vascular tone were the independent variables of the PERMANOVA models, cohort as the dependent variable, with permutations (n=999) blocked by hemodynamic profile (1-3).

#### Validation part 3: comparison of hemodynamic profiles in the validation cohort with the derivation and control cohorts

Comparisons of VC profiling with the DC and CC profiles were first made qualitatively by superimposing scatter plots (cardiac index vs. vascular tone) of the cohorts, along with their hemodynamic profiles. Quantitative analyses were performed by PERMANOVA, in which VC profiling was compared to the DC and CC profiles for multivariable similarity of group composition by cardiac index and vascular tone. In each of two PERMANOVA models (VC vs. DC, VC vs. CC), observations/patients in the comparator cohort (VC) were combined with the reference cohort (DC or CC) into a single dataset with the following variables for all observations: cardiac index, vascular tone, hemodynamic profile number assigned during clustering, and cohort name. Cardiac index and vascular tone were the independent variables of the PERMANOVA models, cohort as the dependent variable, with permutations (n=999) blocked by hemodynamic profile (1-3). We chose α = 0.30 as the level of statistical significance for the hypotheses that cardiac index and vascular tone within each profile differed between cohorts (H_1_: VC vs. DC, H_2_: VC vs. CC). R^2^ for each model were reported, representing the proportion of between-cohort variance in cardiac index and vascular tone within each profile 1-3.

#### Comparison of validation cohort profiles by clinical features

In the DC^18^, no significant (α=0.05) difference in clinical variables was detected, suggesting that the hemodynamic profiles were not readily explained by other common clinical markers and therefore more likely to be a novel marker unto themselves. In the VC, patients were compared by profile for each clinical variable and CDR similarly. Continuous variables were compared with the non-parametric Kruskall-Wallis test or parametric ANOVA. Categorical variables were assessed with the chi-square test.

#### Comparison of validation cohort profiles with clinical study outcomes

The primary and secondary outcomes were assessed first by survival analysis. Kaplan-Meier curves stratified by hemodynamic profile were produced and then compared with the log-rank test. Survival for the secondary outcome was defined as freedom from any fatal or non-fatal adverse event in the hierarchy. Odds ratios (OR) with 95% confidence intervals (CI) were calculated for between-profile comparisons of the categorical parameterization of the primary (binary) and secondary (ordinal/ranked) outcomes.

#### Sensitivity analysis 1: Validation cohort hemodynamic profile as unique novel parameter versus being a simple combination of conventional clinical features

Principal components analysis (PCA), stratified by VC hemodynamic profile, was performed including all variables collected. If between-profile differences could be easily explained by more conventional clinical variables, then relatively few variables should account for a large amount (>75%) of between-profile variance. We reasoned that clinicians likely could not intuit the same information provided by the profiles if it required >3-4 variables to explain the single profile variable.

#### Sensitivity analysis 2: differences between validation cohort profiles in clinical markers of instability and/or severity

VC hemodynamic profiles were compared by the observed (actual) ED discharge rate in the sample, and by clinical features which would suggest to the EP an obvious need for hospital admission (i.e. potential indications of a patient not being safe for ED discharge). Based on local practice patterns, we considered a clear indication for admission as any critical care intervention in the ED (positive pressure ventilation (PPV), inotropes, IV vasodilators, IV rate control for tachydysrhythmia) or unstable vital signs (respiratory rate > 30 or < 6, heart rate >120 or < 50, SBP >200 or < 90, SpO2 < 88% or requiring supplemental oxygen).

## Results

### Participants and description of the validation cohort

Of 351 patients screened against inclusion criteria for the VC and who consented for hemodynamic monitoring, 17 did not begin monitoring in the ED, 4 did not have a recorded EF, and 78 did not have AHF on diagnostic adjudication (Figure 1). The 257 remaining patients had a median age of 60 years (interquartile range{IQR} 49-67) and 45% were female.

**Fig 1.**
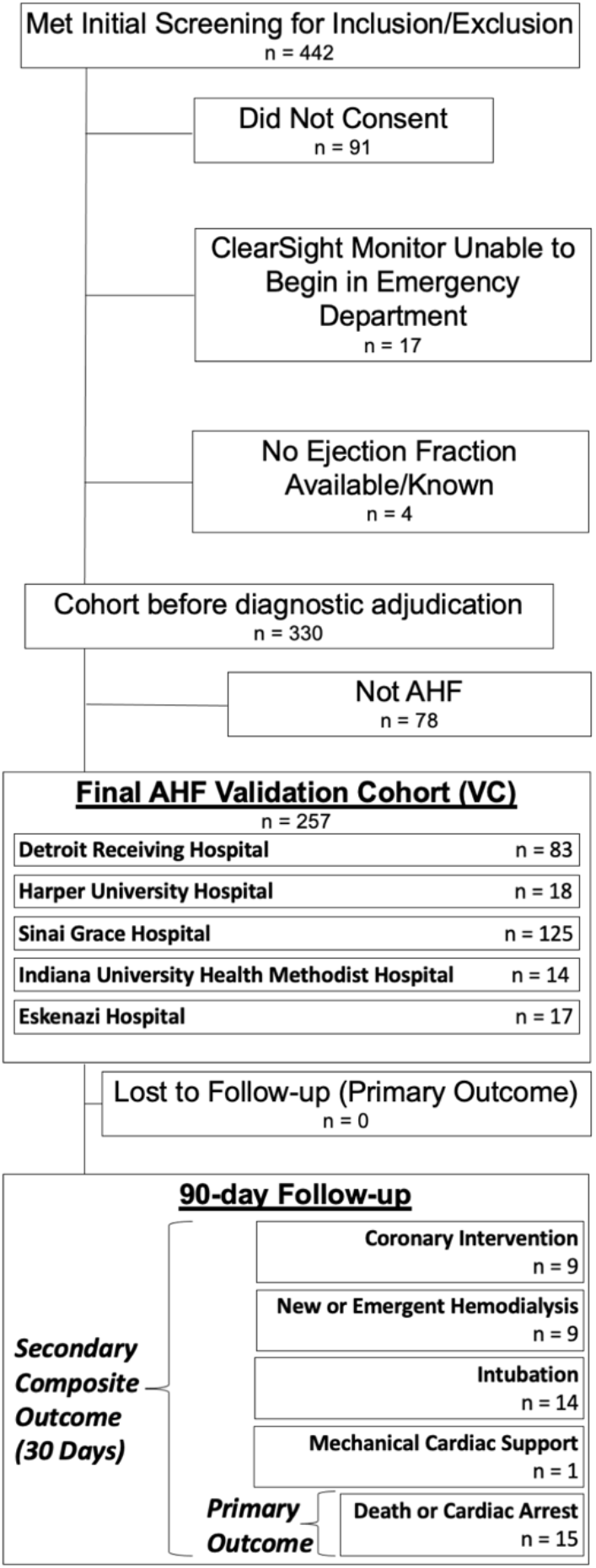
Validation Cohort Flow Diagram. AHF = Acute Heart Failure.

Table 1 presents VC patient demographics, medical history, outpatient medications, initial vital signs, labs, interventions, clinical testing, risk scores, and initial values for cardiac index and vascular tone. 25% required supplemental oxygen, and 9% received a critical care intervention. 47% had HF with reduced EF (HFrEF), 79% of whom were adherent to guideline directed medical therapy. Median (IQR) for cardiac index and vascular tone were 2.40 L/min/m^2^ (2.00-3.08) and 3196 dynes-sec/cm5/m2 (2578-3919), respectively. Characteristics of the DC have been described previously^18^.

**Table 1.**
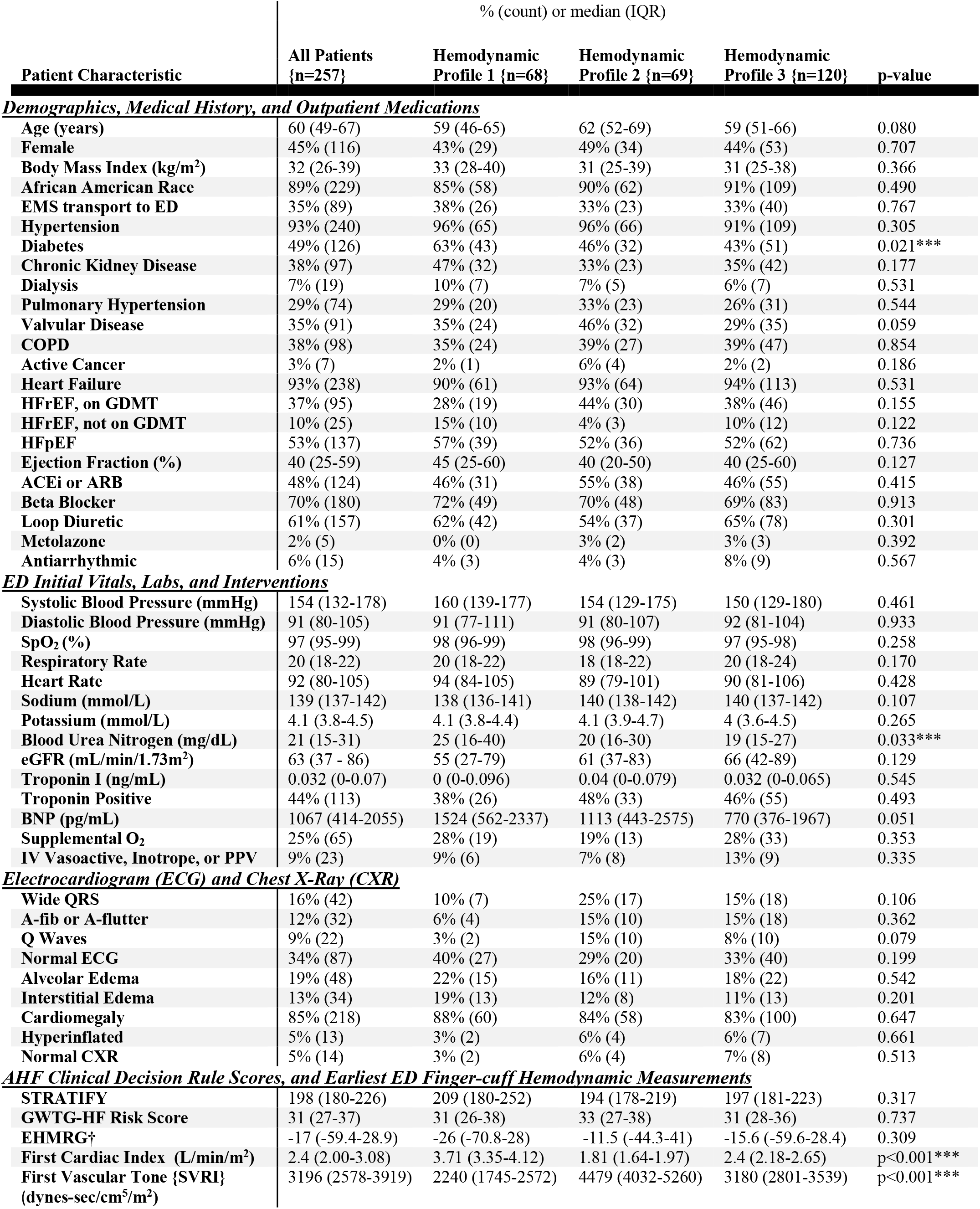
Clinical Characteristics of Patients in the Validation Cohort. EMS = Emergency medical services; ED = Emergency department; COPD = Chronic 405 obstructive pulmonary disease; HFrEF = Heart failure with reduced ejection fraction; HFpEF = HF with preserved EF; ACEi = Angiotensin converting enzyme inhibitor; ARB = Angiotensin receptor blocker; SpO2 = Oxygen saturation; eGFR = estimated glomerular filtration rate; BNP = Brain natriuretic peptide; O2 = oxygen; GWTG-HF = get-with-the guidelines heart failure; EHMRG = Emergency Heart Failure Mortality Risk Grade. †EHMRG as calculated in Table 1 excludes patients who were dialysis dependent, as the EHMRG was derived and validated in a population excluding such patients. The total cohort and profile sizes without dialysis history were total cohort n=238, profile 1 n=61, profile 2 n=64, profile 3 n=113. None of the patients with dialysis history died within 90 days.

### Hemodynamic profiling in the validation cohort

The consensus clustering algorithm was performed in the VC based on cardiac index and vascular tone (SVRI). Inspection of the consensus dendrograms^40^ (Supplemental Figure S2) for each k clusters 1-10 showed the cleanest divisions to occur when the data was divided into k = 3-4 groups. Inspection of the delta area change in consensus score CDF^40^ for K 1-10 (elbow plot, Supplemental Figure S3) show minimal improvement in area under the CDF curve for k>3.

Taken together, these suggest that further divisions of the data (i.e. more profiles / increasing K) beyond k=3 resulted primarily in data sorting at random rather than improved classification^40^ by cardiac index and vascular tone in the VC. Consequently, the clustering and internally-validation performed by the consensus algorithm at k=3 were designated hemodynamic profile 1 (n=68), 2 (n=69), and 3 (n=120) in the VC.

Cardiac index, vascular tone, and clinical characteristics by profile are presented in Table 1. Profiles 1-3 resembled each other (p>0.05) for all clinical variables except BUN and history of diabetes, with each being highest in profile 1 and lowest in profile 3 (Table 1). The 3 validated clinical risk scores (EHMRG, GWTG-HF, and STRATIFY) had no statistically significant difference between profiles (Table 1).

### Comparison of hemodynamic profile composition in the validation cohort to the derivation and control cohorts

Figure 2 shows cardiac index and vascular tone for the 3 profiles in the VC alone (panel A) and compared to the DC^18^ (panel B) and CC (panel C) patients’ hemodynamic profiles. Multivariate statistical comparison by PERMANOVA did not show the profiles in the DC^18^ and VC to be significantly different at the prespecified α=0.3 threshold (Figure 2B, p = 0.59, R^2^=0.016). A significant difference (PERMANOVA p=0.001, R^2^=0.159) in cardiac index and vascular tone was present between the VC and CC profiles (Figure 2C).

**Fig 2.**
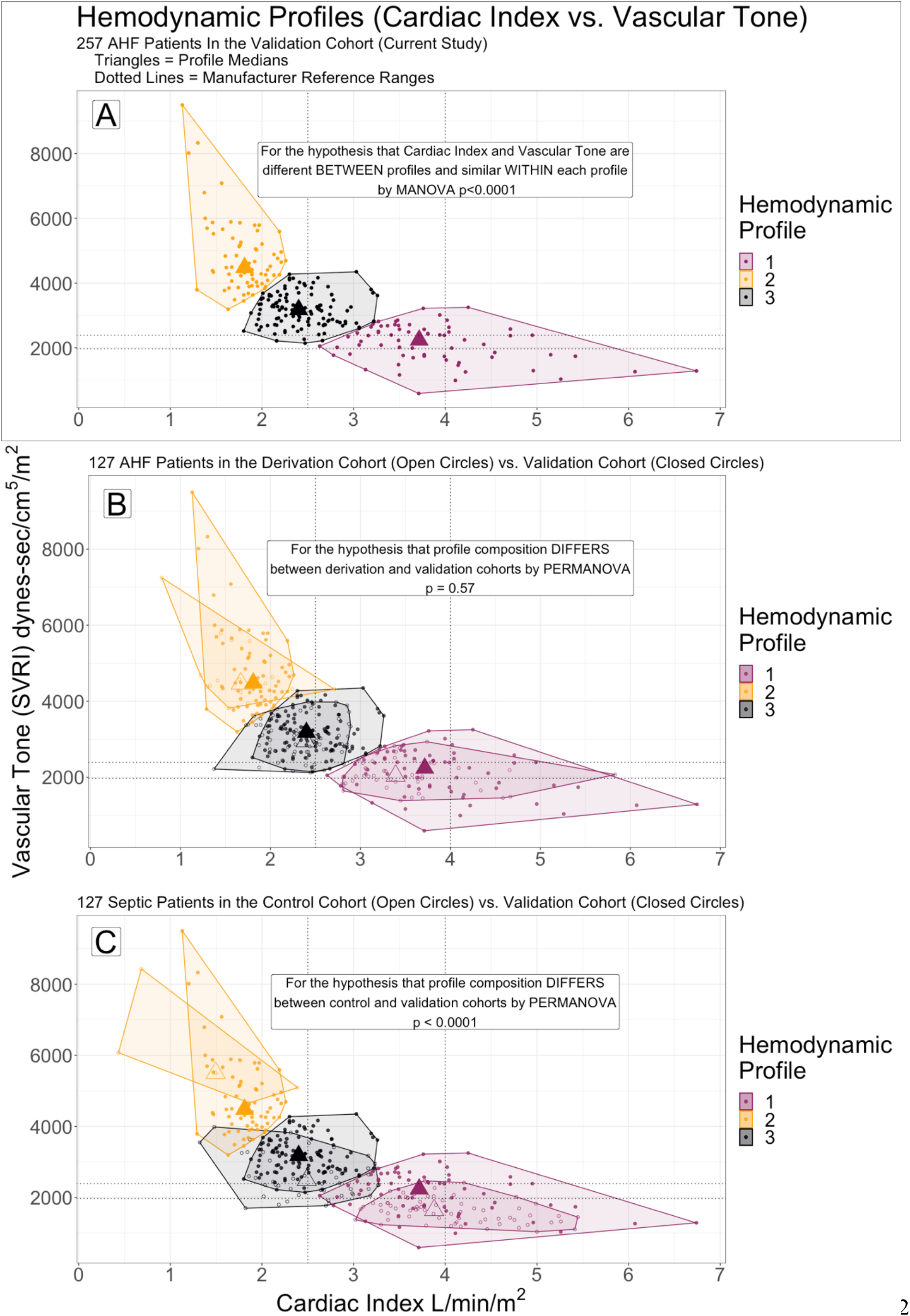
Hemodynamic profiles by cardiac index vs. vascular tone (systemic vascular resistance index, SVRI) Profiles were numbered similarly to facilitate between cohort comparisons: 1 (purple) - lowest SVRI and highest cardiac index, 2 (gold) - highest SVRI and lowest cardiac index, 3 (black) cardiac index and SVRI between profiles 1 and 2. (A) Profiling in the validation cohort (VC) alone. (B) The VC patients and their profiles are overlayed with the derivation cohort (DC). Patients in the DC had acute heart failure and were monitored in the emergency department like the VC, but were enrolled in a prior study (external cohort). Few patients classified in a particular profile in the VC would have been classified differently in the DC. (C) The VC overlayed with the control cohort (CC). The CC included patients enrolled in the same study as the DC, but who had sepsis rather than AHF. Profiling in the CC differed from VC, with several VC patients who would have been classified in a different profile by profiling of the CC (and visa versa).

### Clinical outcomes by hemodynamic profile in the validation cohort

Outcomes rates by profile in the VC are presented in Table 2. 89% of patients were admitted to the hospital or an observation unit, 6% died within 90 days (primary outcome), and 7% experienced ≥1 fatal or non-fatal 30-day adverse event in the composite secondary outcome (Table 2). 90-day mortality (primary outcome) was significantly more likely (OR = 5.0, 95%CI 1.4-18.0) in Profile 3 compared to Profiles 1 or 2 (Table 2). Profile 3 also had shorter time to death than 1 or 2 (Table 2), including every death within 30 (p=0.049) and 60 days (p<0.001).

| Outcome |  | % (count) or mean (SD) |  |  |  | p-value |
| --- | --- | --- | --- | --- | --- | --- |
|  |  | All Patients {n=257} | Hemodynamic Profile 1 {n=68} | Hemodynamic Profile 2 {n=69} | Hemodynamic Profile 3 {n=120} |  |
| <u>Mortality (Primary Outcome)</u> |  |  |  |  |  |  |
|  | 30-Day | 2% (6) | 0% (0) | 0% (0) | 4% (5) | p=0.049*** |
|  | 60-Day | 4% (11) | 0% (0) | 0% (0) | 9% (11) | p<0.001*** |
|  | 90-Day | 6% (15) | 3% (2) | 1% (1) | 10% (12) | p=0.027*** |
|  | Time to Death (days) | 87.3 (12.6) | 89.7 (1.7) | 89.7 (2.2) | 84.6 (17.9) | p=0.016*** |
| <u>30-Day Ranked (0-5) Composite of Adverse Events (Secondary Outcome)</u> - adapted from Collins 2015<sup>31</sup> |  |  |  |  |  |  |
| 0 | Event-free at 30 days | 93% (239) | 87% (59) | 99% (68) | 84% (101) | p=0.008*** |
| 1 | Invasive Cardiac Procedure | 5% (12) | 6% (4) | 1% (1) | 6% (7) | p=0.187 |
| 2 | New or Emergent Dialysis | 3% (8) | 3% (2) | 0% (0) | 5% (6) | p=0.300 |
| 3 | Intubation | 5% (13) | 6% (4) | 0% (0) | 8% (9) | p=0.181 |
| 4 | Mechanical Cardiac Support or Transplant | < 1% (1) | 0% (0) | 1% (1) | 0% (0) | p=0.601 |
| 5 | Death or Cardiopulmonary Resuscitation | 2% (6) | 0% (0) | 0% (0) | 4% (5) | p=0.049*** |
|  | Mean Rank of Worst Event in 30 days (0-5) | 0.30 (0.98) | 0.28 (0.79) | 0.06 (0.48) | 0.46 (1.24) | p=0.003*** |
| <u>Other Outcomes</u> - all at 30-day follow-up unless otherwise indicated |  |  |  |  |  |  |
|  | Discharged from Emergency Department | 11% (28) | 10% (7) | 14% (10) | 9% (11) | p=0.268 |
|  | Admission or Transfer to ICU | 18% (45) | 22% (15) | 15% (10) | 17% (20) | p=0.480 |
|  | Index Hospitalization Length of Stay (days) | 4 (4) | 5 (5) | 4 (3) | 4 (4) | p=0.935 |
|  | ED Revisits for AHF | 18% (46) | 18% (12) | 15% (10) | 20% (24) | p=0.635 |
|  | ED Revisits, All-Cause | 26% (68) | 31% (21) | 22% (15) | 27% (32) | p=0.478 |
|  | AHF Readmissions | 16% (41) | 16% (11) | 13% (9) | 18% (21) | p=0.722 |
|  | All-Cause Readmissions | 22% (56) | 27% (18) | 17% (12) | 22% (26) | p=0.436 |
|  | ICU or Death | 18% (46) | 22% (15) | 15% (10) | 18% (21) | p=0.507 |

Comparison of 30-day events by profile are presented in Figure 3A and Table 2. Figure 3B shows rates for ED critical care interventions, unstable vital signs, dispositions, and loop diuretic administration all of which were similar between profiles (p>0.05). A 30-day fatal or non-fatal adverse event (secondary outcome) occurred in 16% of profile 3 patients, 13% profile 1, and 1% of profile 2 (p=0.008). Median event severity/rank in profile 2 was lower compared to 1 or 3 (p=0.003, Table 2). The likelihood of any 30-day adverse event (Table 2) was higher in Profile 3 vs. Profile 2 (OR=12.8, 95%CI: 1.7-97.9) and Profile 1 vs. 2 (OR=10.0, 95%CI: 1.2-81.2), but similar in Profile 3 vs. Profile 1 (OR=1.28, 95%CI 0.5-3.0).

**Fig 3.**
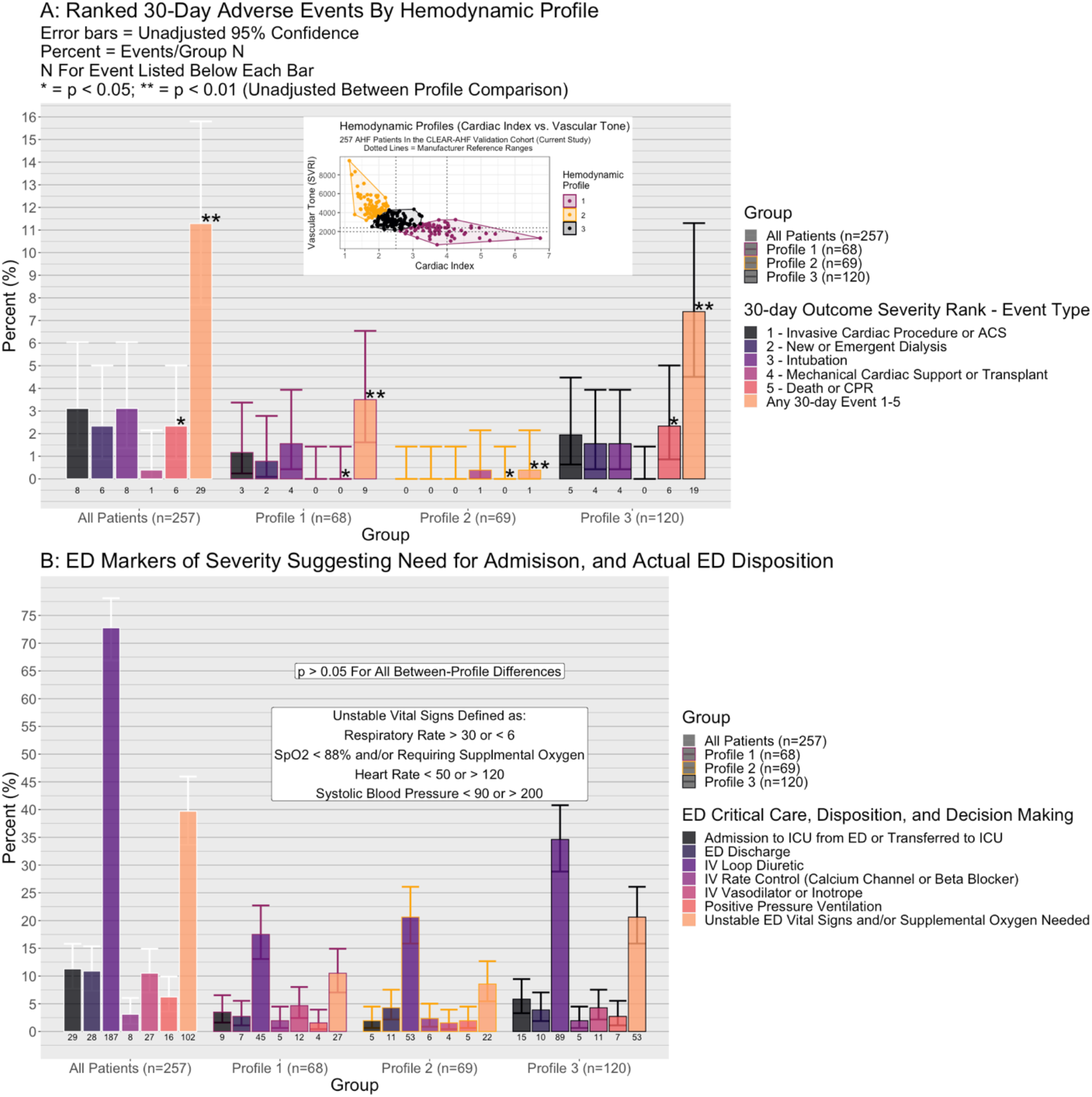
Comparison of cardiac index vs. vascular tone hemodynamic profiles by 30-day adverse events, emergency department (ED) characteristics, and ED disposition. (A) Hemodynamic profiles 1-3 in the validation cohort (inset) are compared by individual components of the composite 30-day secondary outcome. Compared to profile 2 (gold in inset), profile 3 (black) and profile 1 (purple) had greater rates of any outcome in the composite. (B) The were no statistically significant differences between profiles in actual ED disposition decisions (ICU, or discharge from ED), ED treatments administered, or the presence of unstable vital signs or need for supplemental oxygen.

Figure 4 presents Kaplan-Meier survival for mortality (4A) and the secondary outcome(4B) stratified by profile and compared at each of 30, 60, and 90 days (all log-rank p<0.05, both outcomes). Time-to-death was worst in profile 3 (90-day hazard ratio {HR} = 4.83, 95%CI 1.36-17.1), while time to any event (90-day HR = 0.36, 95%CI: 0.15-0.85) was best in profile 2.

**Fig 4.**
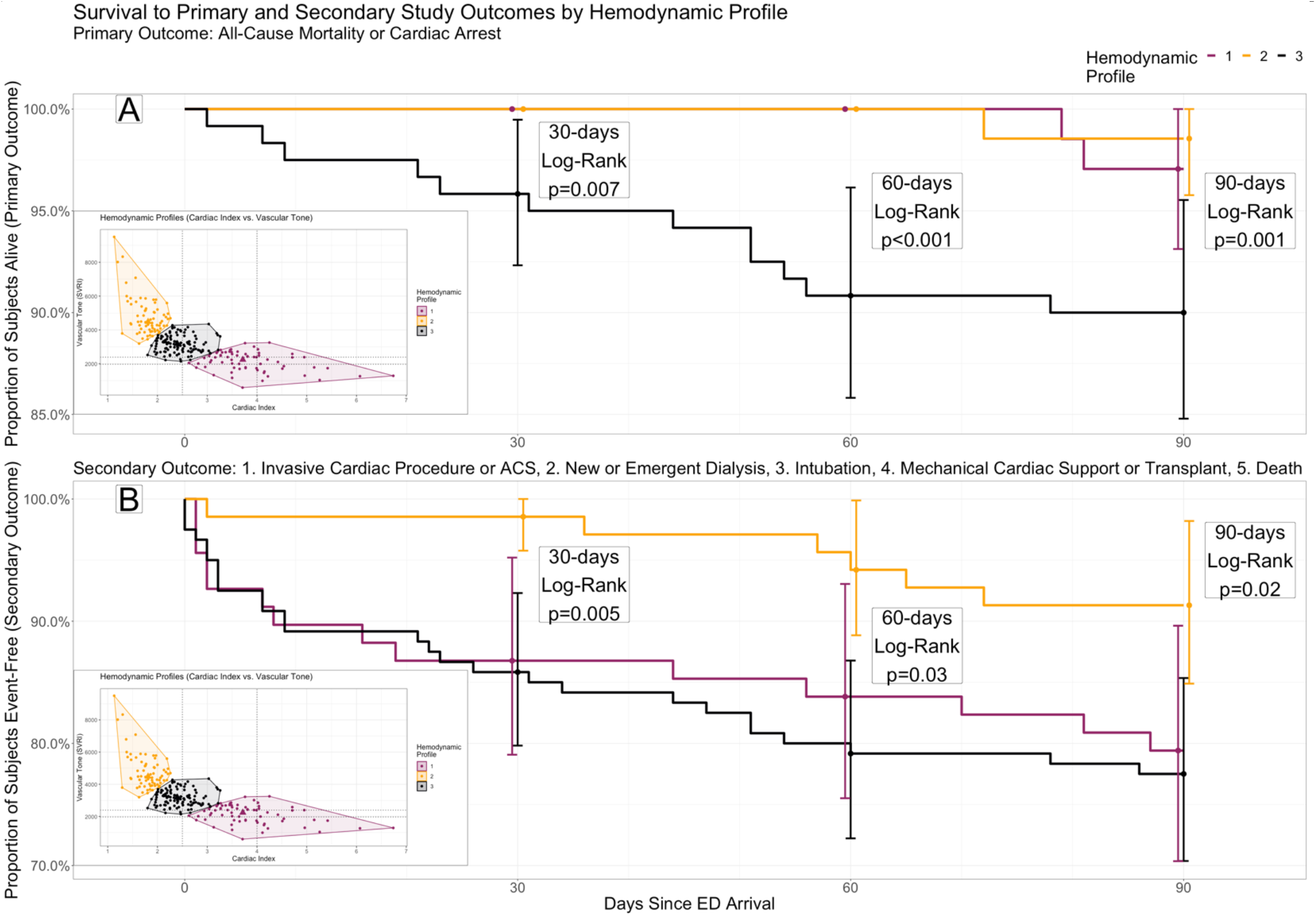
Survival to primary and secondary study outcomes by hemodynamic profile. Kaplan Meier curves for three hemodynamic profiles by cardiac index and vascular tone (see inset) in the validation cohort, through 90-day follow-up. (A) Primary outcome: all-cause mortality or cardiac arrest. Profile 3 (black) has significantly worse survival compared to profiles 1 or 2 (purple and gold, respectively) through each of 30, 60 and 90 days. (B) Secondary outcome: a composite of invasive cardiac procedure, new or emergent dialysis, intubation, mechanical cardiac support or transplant, and death or cardiac arrest. Profile 2 has significantly better event-free survival compared to profiles 1 or 3 through each of 30, 60 and 90 days.

### Sensitivity Analyses

On principle components analysis of the VC hemodynamic profiles, using all clinical variables collected (Table 1) other than cardiac index and vascular tone, we found that ≥7 principal components were required to explain ≥75% of between-profile variance.

116/257 (45.1%) patients met our specified criteria (Methods - Sensitivity analysis 2) for a “clear indication” for hospital admission in the ED. After excluding these patients, just 12.1% of those remaining (17/141) were discharged from the ED, but 56.7% (80/141) were hemodynamic Profile 1 or 2 (low risk for 90-day mortality) and 30.0% (42/141) were Profile 2 (low risk for any 30-day adverse event). If Profile 2 vs. Profile 1 or 3 was used as a discharge criterion in these 141 patients without a clear indication for hospital admission, significantly more patients (p<0.001) would have been discharged from the ED compared to the actual discharge rate, without any missed deaths (100% negative predictive value for Profile 2).

## Discussion

In this multicenter prospective observational study, we externally replicated non-invasive hemodynamic profiling of ED AHF patients by cardiac index and vascular with a finger-cuff monitor. Hemodynamic profiling has long been used to subgroup AHF patients by clinically-important differences in physiology, particularly in the relationship between cardiac index and vascular tone. Despite being recommended as part of routine AHF assessment in guidelines^1,22,23^, invasive measures are so specialized and uncommon^19^ that ED use is virtually unheard of, while current non-invasive methods^20,21^ based on physical exam are subjective and lack sufficient interrater reliability^12,18,24,41^ in the ED. Finger-cuff monitors are a non-invasive approach which provides objective data, and the replication of the exploratory cluster analysis results in the prior study’s DC^18^ adds external validity to the observed profiles as a novel marker in ED AHF patients.

In the prior study^18^ patients had marked heterogeneity by cardiac index and vascular tone and clustered into three novel two-dimensional profiles. We were able to replicate these profiles *de novo* in a prospectively enrolled and diagnostically adjudicated external cohort, whereas the DC was a retrospective analysis without diagnostic adjudication. Additionally, we used a specialized cluster analysis procedure which was less reliant on analyst assumptions and theoretically more robust to data overfitting than what was used in the DC’s profiling. Our findings nevertheless replicated the heterogeneity and distribution of cardiac index and vascular tone first noted in the DC study^18^, while clearly differing from the septic CC. Replication achieved by these methods add rigor and bolster the case that these hemodynamic profiles represent a true feature of AHF patient is the ED, rather than artifact and overfitting of the prior study’s single retrospective sample.

Both 90-day mortality and a composite of 30-day adverse events differed significantly between profiles in the VC. Assessing these differences was a novel and primary goal of the current study, since the DC study was neither powered for nor designed^18^ to test differences in clinical outcomes between profiles. The between-profile differences in adverse events observed were clinically significant: no patients died within the first 60 days outside the high-risk profile 3, and death remained five times more likely in profile 3 at 90 days. Additionally, the low-risk profile 2 had roughly 12 times lower likelihood of any adverse 30-day event compared to profiles 1 or 3.

Novel risk markers for AHF are needed in the ED^1^, and particularly markers of low risk ^1,2,8-10^. Over 80% of the 1 million AHF patients presenting to US EDs annually are admitted, including over 90% of the current sample, many of whom are at low risk for short term adverse events^1^. The burden on patients and healthcare resources is correspondingly high. Among the 141 patients in the VC who lacked one or more clear criteria for ED-to-hospital admission at our institutions (sensitivity analysis 2), the low-risk profile 2 was present in more than double the actual number of patients discharged. Similarly, over 57% of those without clear admission criteria were in profile 1 or 2, among which no patient died within 60 days. Physicians were blinded to monitoring of cardiac index and vascular tone in this study, and it is possible that knowledge of a patient’s hemodynamic profile would have improved risk-stratification and facilitated ED discharges.

As in the DC study^18^, hemodynamic profile was not clearly explained by other clinical variables. Two validated CDRs related to the primary outcome, and one for the secondary outcome, did not differ significantly between the high vs. low-risk profiles observed. Among a long list of common clinical variables, only BUN and history of diabetes differed between profiles. While the absence of diabetes and a lower BUN would be expected to correlate with better clinical outcomes, these variables were paradoxically the lowest in the highest risk hemodynamic profile (profile 3). Principle components analysis (sensitivity analysis 1) failed to yield any simple combination of clinical variables which would explain the variance in patients’ hemodynamic profile. It is unlikely, based on the results in the VC and the prior description of the DC^18^, that these hemodynamic profiles are simple functions of more common and available clinical measures. Instead, these results suggest that hemodynamic profiling by finger-cuff monitor cardiac index and vascular tone add novel information not already captured in the standard of care. Further research is needed to assess if the information added, particularly regarding association of these profiles and clinical outcomes as a risk-measure, is incremental with other established AHD risk measures. A pre-planned analysis of the VC to assess for incremental value in risk-stratification and prognosis is currently underway.

## Limitations

This study had several limitations. First, finger-cuff hemodynamic monitors have a roughly +/- 30% error in cardiac index compared to invasive monitoring^18,35^, which is below the level of construct validity to completely replace invasive catheter based methods in the cardiac ICU^18,35^. Nevertheless, invasive monitoring is not feasible in the ED and physical exam based non-invasive alternatives lack reliability and accuracy^12,18,24,41^, making the error rate in finger-cuff monitors likely the best achievable in this patient population and setting at present. Second, in our approach to replication and validation using PERMANOVA means we failed to reject the hypothesis that the VC did not replicate the DC profiles, which is not the same as accepting the hypothesis that they were the same. We used a more conservative alpha threshold of 0.3 to test this hypothesis to decrease the chance of type II error, but we cannot guarantee that additional replication studies or a larger statistical power would have failed to reject the null hypothesis. Moreover, the clear differences of the VC and CC profiles enhance our confidence in the results, given that the CC patients were enrolled at the same times and hospitals as the DC but with a different underlying ED diagnosis (sepsis, rather than AHF). Third, a 90-day primary outcome may be a longer follow-up than some EPs would deem clinically relevant. However, there were statistically significant between-profile differences at 30 and 60 days for mortality, and at 30-days for all adverse events. Fourth, our study was performed at 5 high-volume academic EDs, and results may not generalize to dissimilar settings or AHF patient populations significantly different than our sample (Table 1). Fifth, unaccounted for lost-to-follow-up is possible for the secondary outcome, such as if a patient had an adverse event at an outside hospital. We confirmed 100% of patient follow-up for the primary outcome at 90 days between the use of telephone follow-up, HIEs including the largest hospital systems near the study sites, and dual-review with adjudication for outcome record review. Finally, while no clear combination of individual variables or CDRs appear to explain the difference in profiles, this does not imply that adding the hemodynamic profiles would add incremental prognostic value to existing AHF risk measures. Rather, incremental prognostic utility is a separate question, to be addressed in a pre-planned future analysis of the VC.

## Conclusion

In this prospective observational cohort study, we validate 3 distinct hemodynamic profiles of ED AHF patients by cardiac index and vascular tone, as measured on a non-invasive finger cuff monitor and described in prior work^18^. Mortality and a composite of adverse short-term events differed markedly between these profiles, suggesting a potential for use in the ED as a marker for risk-stratification.

## Data Availability

Data produced in the present work, and code for reproduction of the results in R, are contained in peer review submission. These will be made available upon publication, or reasonable request.

## Supplemental Material

### S1: Further methodological detail on consensus clustering, and contrast to k-means cluster analysis

The prior study used k-means clustering to derive three hemodynamic profiles among the PREMIUM registry’s AHF patients (derivation cohort, DC). K-means clustering has two major weaknesses: 1. the data analyst must decide before clustering how many clusters (k) to subgroup the data by, and 2. the clustering is a direct function of the data and therefore does not guarantee clusters can be replicated in an external sample. Both weaknesses have the potential to lead to model overfitting. Consensus clustering is a machine learning technique which improves on k-means cluster analysis primarily by addressing these two weaknesses, and thereby reducing the chance of an overfit model^18^. First, the technique does not assume 3 clusters (or any other number) to be the number of truly distinct hemodynamic profiles present. Rather than being pre-specified, the number of distinct hemodynamic profiles to group observations into is iteratively tested for every possible number of clusters 1-10. Second, clustering is replicated 1000 times on random sampling of the dataset after introducing random perturbations of data points, which decreases the potential for bias related to the sampling of the cohort itself. At each number of clusters tested (1-10), the resampled cluster analyses are pooled to arrive to a “consensus” score wherein patients are grouped to maximize similarity in cardiac index and vascular tone within a cluster, while maximizing differences for patients in different clusters. An elbow plot of the delta area change in cumulative distribution function and other diagnostics are then produced which indicate the point at which splitting the data into additional clusters no longer enhances within-profile similarity or between profile differences for CI and SVRI (i.e. the point past which further subdivisions/higher K reclassifies data by random/meaningless divisions). For further methodological details on consensus clustering, see Wilkerson et al.^40^. For consensus clustering in the validation cohort (VC) for this study, cardiac index and vascular tone were log transformed and scaled to facilitate efficient clustering by Euclidean distance.

### S2: Consensus Clustering Dendrograms

Consensus Clustering^40^ expands on K-means cluster analysis in two primary ways: 1. Repeating clustering with random perturbations of the data to internally-validate cluster stability, and 2. Assessing clustering at multiple levels k (k = number of times the data is divided). Dendrograms below, for each k tested in the dataset (2-10), show the proportion of repeated clustering runs in which each segment of the data was assigned to a given cluster. Dark blue indicates a data segment which is always assigned to the same cluster (i.e. maximum consensus/cluster stability) while lighter blues indicate where a data segment was fit into one of multiple clusters based on the random data perturbations (low consensus/cluster stability). A lack of consensus/cluster stability suggests that clustering is more due to random chance than true similarity within the data clusters.

In the dendrograms (this page and next), cluster stability appears to reach a maximum at k=3, with nearly random clustering for k>5. Thus, a choice of 3 groups (3 divisions of the data) appears to maximize within cluster similarity and maximize between-cluster differences.

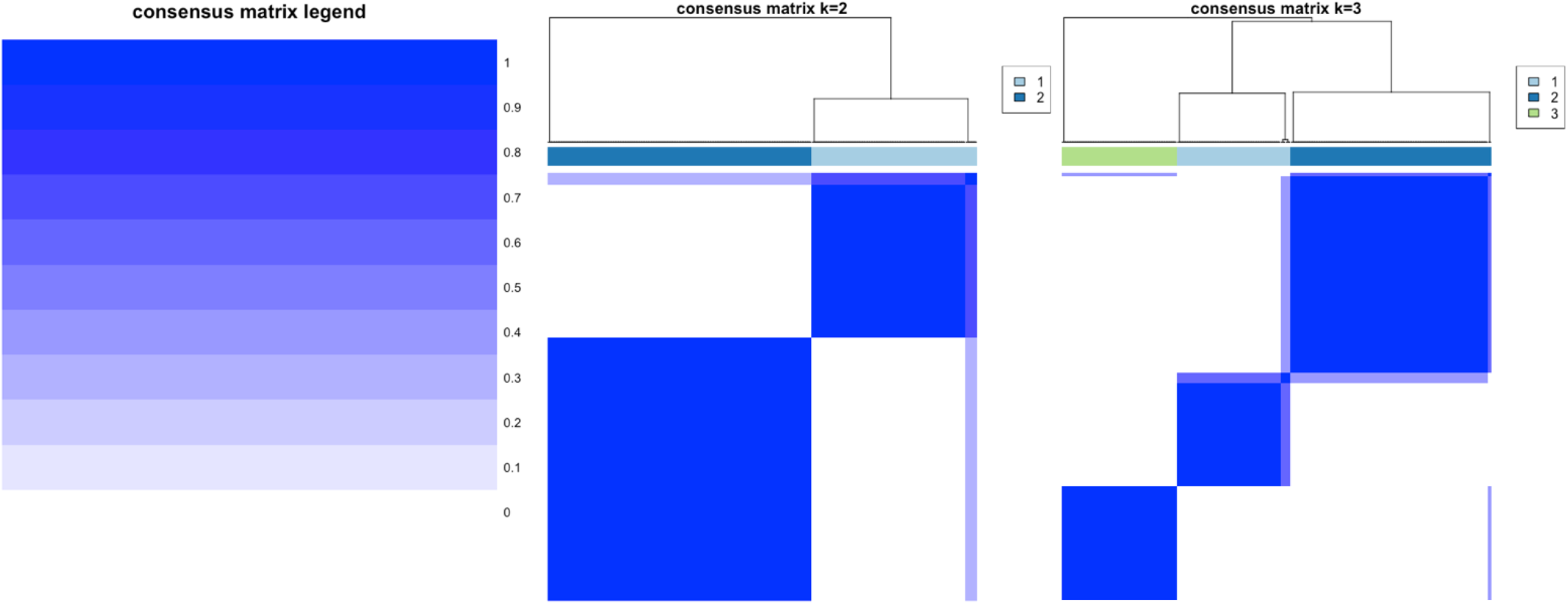

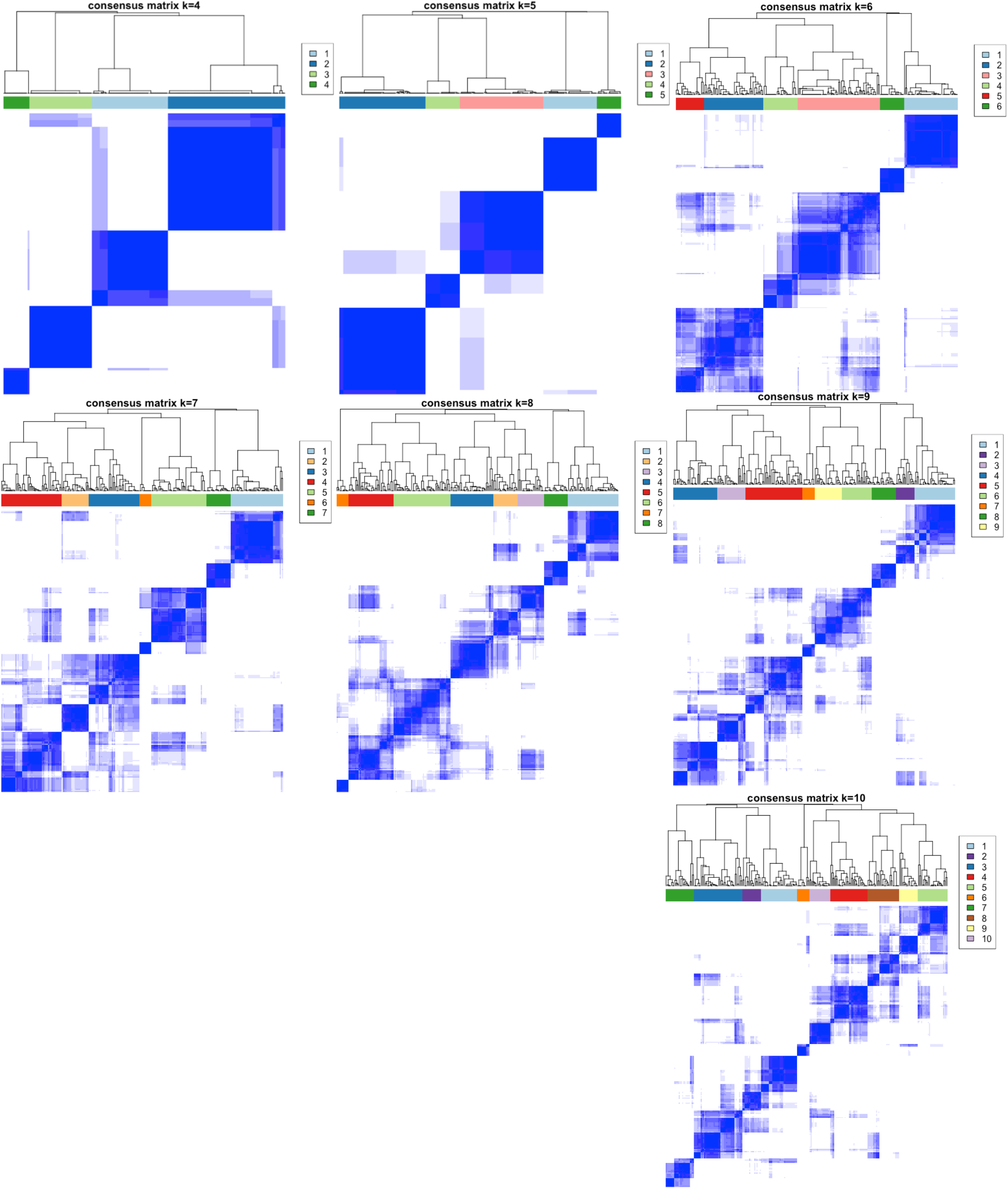

### S3: Delta-area under the cumulative distribution function (CDF) for each additional level K in consensus clustering

In this elbow plot, the relative change in consensus CDF is shown for each additional data division (i.e. 1 increment increase in k) during consensus clustering. The elbow, where further increases in k yield relatively little change in area under the CDF curve, indicates the K at which further additional clusters add little to the goal of class discovery. The elbow occurs around k=4 in the validation cohort. This further supports what is suggested in supplemental figure S2: additional divisions of the data beyond k=3 yields little improvement in class discovery and stability.

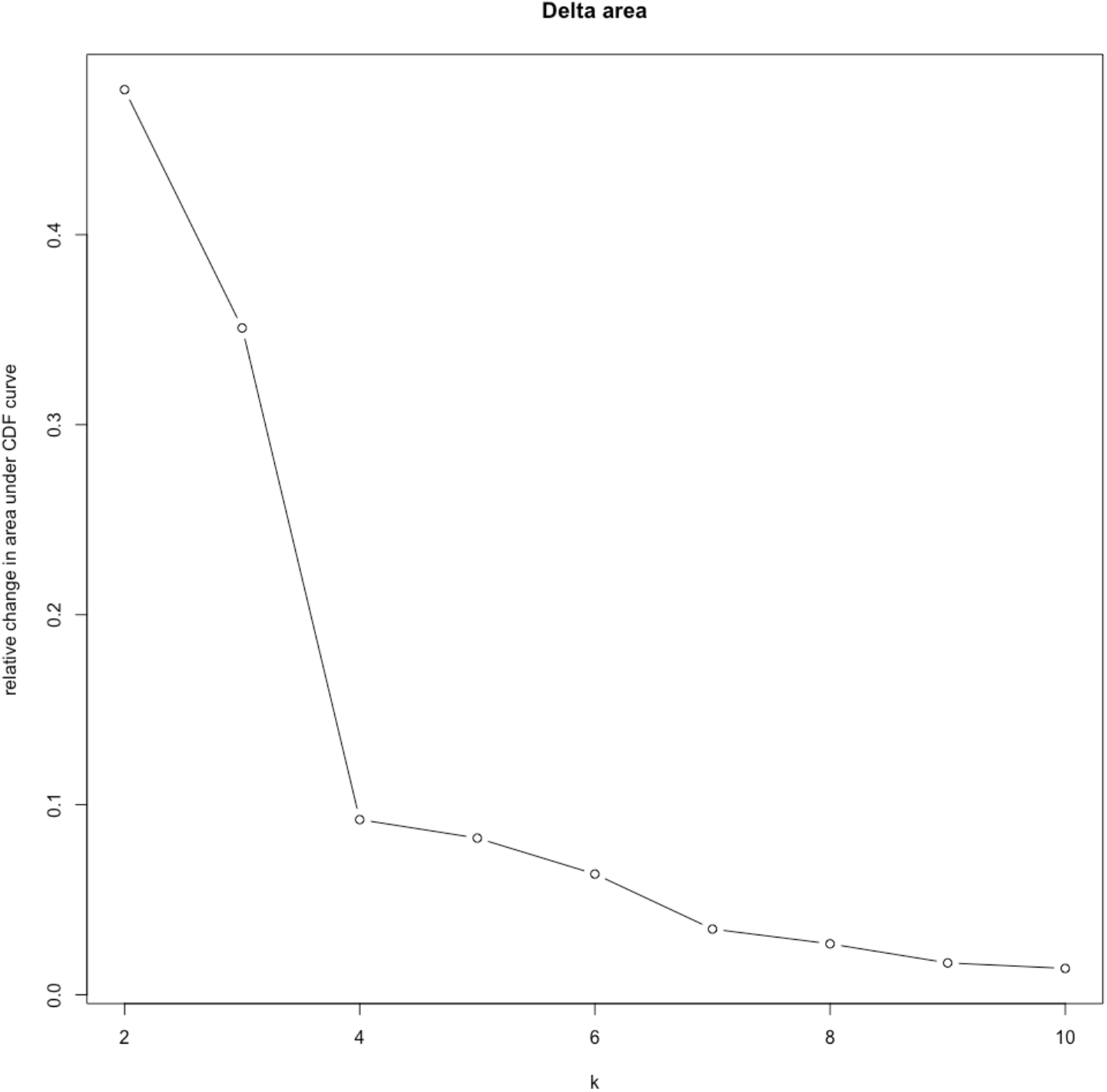

### S4: Code for Analysis and Minimal Dataset

Code for the R statistical programming language, and a minimal data set, to reproduce our results may be found attached in this submission and authorized for public use.

## Acknowledgments

None

